# Financial Feasibility Study and policy recommendations of Incremental Modified Drugs Development by the Domestic Pharmaceutical Industry

**DOI:** 10.1101/2024.07.29.24311184

**Authors:** Manthana Laichapis, Rungpetch Sakulbumrungsil, Khunjira Udomaksorn, Nusaraporn Kessomboon, Osot Nerapusee, Charkkrit Hongthong, Sitanun Poonpolsub

## Abstract

The Thai pharmaceutical industry aims to strengthen its drug system in accordance with the National Strategic Master Plan, emphasizing sustainable development, particularly in biologics and herbal products, to achieve self-reliance. Current efforts are mainly focused on generic drug production, but there’s a significant need for Research and Development (R&D) in Innovative Medicines (IMDs). This study explores the financial feasibility of locally developing an IMDs dosage form. To assess this feasibility, a mixed-methods approach was used, incorporating a literature review, surveys, and interviews. This process involved selecting types of IMDs, constructing financial models, determining cost structures, and conducting a thorough feasibility analysis. The results indicated that a sustained-release dosage form was the most viable option. The analysis took into account total development costs, payback periods, growth rates, and the revenue required to recoup investments. It was found that IMD development is associated with higher costs and longer durations compared to new generic drugs.The study identified several challenges, such as the high cost of clinical studies, extended development times, market feasibility, and drug selection difficulties. Policy recommendations were made to address these challenges, including incentives for clinical studies and fostering industry expertise through collaborative efforts and supportive government policies.In conclusion, the financial feasibility of developing IMDs requires strategic policies and collaboration to overcome these challenges and ensure sustainability. The findings of this study are intended to aid stakeholders in making informed R&D investment decisions.

## Introduction

The Thai pharmaceutical industry has focused on development in accordance with the National Strategic Master Plan (2018-2037), specifically targeting the enhancement of the national drug system and the potential growth of pharmaceutical manufacturing within the country (1), The third national drug policy of 2011 and the national drug system development strategy for 2011-2016 have emphasized the sustainable development of the pharmaceutical industry, including biologics and herbal products to achieve self-reliance (2). The primary objectives include the acceleration of capabilities and the elevation of the pharmaceutical industry through research, development, and production of vaccines, drugs, and biologics. Moreover, promoting local pharmaceutical industries and services is crucial to reduce imports and increase exports.

Currently, the Thai pharmaceutical manufacturing industry predominantly focuses on the production of generic drugs, with an average annual approval of 540 generic drugs as well as new generic drugs (averaging 35 approvals per year). However, the number of new drug formulations developed and approved in Thailand remains relatively small, largely because of the unpreparedness of the pharmaceutical manufacturing industry and relevant regulatory agencies, and the lack of explicit guidelines for the registration of new drugs produced in the country. Research and development (R&D) efforts have become more focused, with the Food and Drug Administration (FDA Thailand) defining seven types of new drugs: 1. New chemical entities (NCEs), 2. New indications, 3. New combinations, 4. New delivery systems, 5. New routes of administration, 6. New dosage forms, and 7. New strengths. Types 2-7, often referred to as incrementally modified drugs or IMDs in many countries. Among these, NCEs require significant investments, technology, and personnel, and Thailand currently lacks sufficient capacity. Over the past decade, pharmaceutical manufacturing companies have invested in R&D and adopted new technologies to develop different forms of finished products (3).

To support the pharmaceutical manufacturing industry, it is imperative to conduct R&D on incrementally modified drugs (IMDs) that possess compounds and efficacies similar to the original drugs but exhibit modified properties and characteristics. This can be achieved through the use of advanced technology platforms to promote sustainable self-reliance in the Thai pharmaceutical industry. Consequently, this study aims to explore the financial feasibility of developing a dosage form for IMDs in the local pharmaceutical manufacturing industry. Thus, it provides an investment proposal to support policymaking and investment decision-making from an industrial perspective.

## Method

The present study employed a mixed-methods approach, incorporating literature review, surveys, and interviews as the qualitative part, to explore the construct of financial analysis, which constitutes the quantitative part.

### Literature review

The literature review conducted to examine the different dosage forms used in the production of existing IMDs, as well as the manufacturing processes involved in IMDs, from the upstream process to the downstream process. Data for the model was collected through the literature review and related documents to assess the situation and ability to manufacture IMDs in Thailand. An interview instrument was developed to interview relevant experts.

### Survey

The survey method was used to identify and estimate costs based on the defined cost structures, which were derived from the ‘Impact of Thai-EU Free Trade Agreement (FTA) concerning Intellectual Property Rights on the Pharmaceutical Supply Chain in Thailand’ study (Liangrokapart et al., 2013)

1. The collection forms with pre-defined cost structures were sent to 5 IMDs (Industrial Manufacturing and Development) experts.
2. The experts provided estimated costs and comments to make the cost structure more valid.

### Interview

The interview process involved selecting experts and individuals with expertise in assessing the situation and ability to manufacture IMDs in Thailand. The recruitment period for the study spanned from August 5, 2021, to August 4, 2022. This period indicates the timeframe during which participants were actively recruited and enrolled in the study.

#### A.) Demographic of informants

The sampling method employed for this study was the snowball sampling technique until data saturation was reached. Participants were recruited from 15 local pharmaceutical industry companies, of which 5 were company owners, and the remaining participants were individuals associated with IMDs development.

#### B.) Interview process

1. The researcher sent interview questions to research participants and schedule individual online interviews. Each interview is expected to take approximately 1 hour. The researcher sought permission from participants to record the interview for data analysis purposes (audio recordings will be destroyed at the end of the project).
2. Participants’ agreement to participate in the research and their decision to participate in the online interview were considered as consent. Participants were not required to sign a letter of consent.
3. Participants who complete an online interview received remuneration of 1,000 baht per person.

#### C.) Question guidelines

1. What are the costs associated with the research and development of a dosage form for IMDs?
2. What are the procedures involved in the research and development of a dosage form for IMDs, and what are the associated costs?
3. What are the costs related to manufacturing technology?
4. What are the costs of conducting clinical and non-clinical studies?

The present study employed a mixed-methods approach, incorporating literature review, surveys, and interviews as the qualitative part, to explore the construct of financial analysis, which constitutes the quantitative part. The methodology was adapted to perform the following steps:

#### 1. Selection of IMD types

Sustained release, first modified dosage form, and most preferable types of IMDs were identified based on the results of a feasibility study of incrementally modified drugs developed by the domestic pharmaceutical industry (4, 5).

#### 2. Development of financial models

Suitable investment models were developed to align the specific circumstances and capabilities of the local pharmaceutical industries.

#### 3. Identification of cost structure and cost estimation

The cost structure was determined, and the costs associated with the manufacturing industry’s investment in IMDs were estimated. A cost structure adapted from a study on the Impact of the Thai-EU Free Trade Agreement (FTA) concerning Intellectual Property Rights on the Pharmaceutical Supply Chain in Thailand was applied. This structure encompasses sourcing, R&D at the lab scale, pilot batch and stability studies, non-clinical trials, clinical trials, registration, and process validation batch stages (6). Costs were estimated based on each step of the drug development process until the drug was launched in the market (7, 8).

#### 4. Financial feasibility analysis of IMDs

Assumptions were set, and data related to investment variables were collected to analyse the financial feasibility of IMDs. Sensitivity analysis was conducted to evaluate the impact of variations in cost, duration, and payback period. Additionally, the cost was estimated based on two scenarios, considering the regulatory requirements of IMDs (9). The first scenario involved conducting only Phase I (scenario 1), whereas the second involved conducting a full clinical trial (scenario 2).

## Results

Based on a prediction market analysis conducted during a feasibility study of IMDs development by the domestic pharmaceutical industry, the sustained-release dosage form emerged as the most preferable option (4). Consequently, the financial feasibility of this particular dosage form was analysed, considering the input data and assumptions outlined in Table 1.

**Table 1.** Input data and assumptions for financial model and financial feasibility study.

|  | Details | Source of data |
| --- | --- | --- |
| Cost of sales | 25% of revenues | Literature review (12) |
| Operational expense | 40% of revenues | Literature review (12),<br>interview |
| Discounted rate | Discount rate 3% | Literature review (13) |
| Interested rate | Interest rate for business is 3% | Interview |
| Tax rate | Corporate tax rate is 20% | Literature review (14),<br>interview |
| Expected payback period | Payback period that investors<br>could be accepted is 5-10 years | Interview |

### Model for financial feasibility analysis

This study utilizes a distinct financial viability analysis model that allows investors to determine the payback period and market growth rate based on a business’s capacity and provides valuable information for assessing the financial feasibility of investing in pharmaceutical R&D. The model encompasses two main components: a cost model for researching and developing the drug dosage form and a sales revenue model that calculates the necessary income for capital return within the specified payback period. It offers flexibility for investors to adjust variables, such as the payback period and market growth rate, aiding in the decision-making process for selecting and developing study drugs.

### Financial feasibility analysis of sustained release dosage form

Based on the information presented in Table 2, the R&D process for new sustained-release forms of IMDs was found to take approximately 7 years for conducting only Phase 1 clinical studies, and 11 years for the full clinical trial. By comparison, the development of new generic drugs typically ranges from 25 to 46 months.(6) The longer duration in the case of sustained-release forms of IMDs can be attributed to the development of new formulations, higher failure rates, increased complexity of analysis, and the inclusion of clinical studies involving a new drug.

**Table 2.** The process and cost of developing IMDs in the form of a sustained release by the domestic pharmaceutical industry.

| **Process** | **Details**<br>**(6)** | **Scenario 1**<br>**(Million USD)** | **Scenario 2**<br>**(Million USD)** | **Information Source** |
| --- | --- | --- | --- | --- |
| **Sourcing** | Local manufacturers choose IMD reference drugs based on marketing, user needs, sales, patents, and suitability. | 0.02 | 0.02 | Interview |
| **R&D lab scale** | The R&D department conducts lab-scale studies to develop suitable formulations for sustained release drugs, including analytical method development and determination of finished product specifications (FPS). | 0.06-0.15 | 0.06-0.15 | Interview |
| **Pilot scale** | After successful drug R&D, pilot batch production begins, followed by stability studies to determine shelf-life specifications. Results are reported to FDA. | 0.27-0.66 | 0.27-0.66 | Interview |
| **Clinical study** |  |  |  |  |
| **-Phase I** | Samples from the pilot batches production will be sent to study the effect of food on bio-efficacy through bioequivalence study and evaluating the effect of alcohol on dose dumping. | 0.29 | 0.29 | Literature review<br>,Interview,(9),(15) |
| **-Phase II** | In case the pharmacokinetics of a new drug are clinically significant different from the reference drug, Phase II and III studies of the new drug may be necessary. | - | 4.29 | Literature review<br>,Interview,(9) |
| **-Phase III** |  | - | 12.86 |  |
| **Registration** | Registration of new drug formulas follows ASEAN Harmonization criteria. Application documents include administration data, product information, quality, safety, and efficacy parts. Non-clinical and clinical study data can refer to recommendations and guidelines. | 0.003 | 0.003 | Interview |
| **Process validation batch** | After obtaining FDA registration, drugs can be produced in commercial batches. Process validation and inspection results are submitted for permission to continue production and distribution. | 0.81-1.97 | 0.81-1.97 | Interview |
| **Total** |  | 1.46-3.09 | 18.60-20.23 |  |
| **Duration (years)** |  | 7 | 11 | Interview |

The development of new sustained-release drugs from existing chemical entities for conducting only Phase I incurs fixed costs ranging from 1.46 to 3.09 million USD. In comparison, the cost of developing new generic drugs ranges from 0.19 to 1.13 million USD(6). Higher costs associated with sustained-release drugs can be attributed to these factors. A significant portion of the drug development costs in this scenario was allocated to process validation batches, accounting for approximately 60% of the total R&D cost. This final step before commercialization requires three consecutive production cycles, leading to substantial capital requirements that can vary based on the complexity and ease of production.

In scenario with full clinical trial, which involves studying the drug from Phase 1 to Phase 3 clinical trials to establish efficacy and safety, the fixed costs for new drug development range from 18.60 to 20.23 million USD. Most of the cost and time in this scenario are allocated to clinical trial studies, constituting approximately 70% of the total cost. Clinical trials are crucial to prove the efficacy and safety of new drugs. Specific processes, sample sizes, and drug types can influence R&D costs.

Investments in the development of new drugs from existing chemical entities incur high costs. The feasibility analysis considers the costs of drug formulation development and expresses them as the income that entrepreneurs should generate to achieve the capitalization point. The feasibility results indicate that, for the base case analysis with an expected payback period of 5 years, the income required to cover the invested capital ranges from 0.20 to 1.80 million USD for the scenario conducting only Phase I, and 3.01 to 27.11 million USD for the full clinical trial scenario (Table 3).

**Table 3.** The feasibility study results of this study show in terms of income that investors should be able to achieve in order to achieve capital gains.

|  | Scenario 1 (Million USD) | Scenario 2 (Million USD) |
| --- | --- | --- |
| Payback period |  |  |
| 5 years payback period | 0.20-1.80* | 3.01-27.11** |
| 10 years payback period | 0.04-1.20 | 0.63-17.87 |
| Duration of research and development |  |  |
| Scenario 1 |  |  |
| 5 years | 0.18-1.66 |  |
| 10 years | 0.26-2.33 |  |
| Scenario 2 |  |  |
| 10 years | 2.85-25.63 |  |
| 15 years | 4.19-37.73 |  |

\* The parameters of scenario 1 base case analysis are that the duration of research and development is 7 years, and the research and development cost is 1.46 million USD.
\*\* The parameters of scenario 2 base case analysis are that the duration of research and development is 11 years, and the research and development cost is 18.60 million USD

Furthermore, with a longer duration of R&D in both scenarios, the annual income that investors must generate will be higher compared with shorter time periods, indicating a higher level of investment. Therefore, if entrepreneurs can expedite the R&D process, they can save investment costs and increase the feasibility of developing these drugs. In cases where formulation and analytical method development carry a high risk of failure and involve numerous areas of development, including more sophisticated clinical studies, investing in R&D processes may require higher capital investment, and investors need to generate higher income to achieve a higher capitalization point.

Considering the payback period for investors, longer payback periods tend to result in a lower capitalization point, requiring lower annual income generation by operators, compared to shorter payback periods. If a drug has a longer lifecycle and does not require frequent development, such as drugs for chronic diseases, a longer acceptable payback period may be applicable.(10) Various factors, including the annual sales growth rate, marketability upon launch, number of competitors, and government policies, influence feasibility assessment.

## Discussion

This study conducted a financial feasibility analysis, specifically focusing on sustained-release dosage forms, which differ from other financial models. The analysis estimated four key constructs: the total cost of IMD development, expected payback period, expected growth rate, and expected revenue that investors could potentially earn to cover the break-even point or generate profit. However, these dosage forms may vary in their active ingredients, making it challenging to estimate sales revenue and market growth without precise information regarding the specific active ingredients being developed for the studied drug form. The methodology chosen in this study is suitable for analyzing the financial aspects of new drugs or innovations despite limited data availability. To strengthen the reliability of the limited data, the triangulation method was employed, enhancing the validity and credibility of the findings.

The findings from each scenario indicate that investments in IMDs are relatively high and require a longer development duration than new generic drugs. A major portion of these investments is attributed to conducting clinical studies for registration, drug selection, and the sufficiency of presented data to confirm efficacy and safety. Financial feasibility considerations are also influenced by an entrepreneur’s capacity to invest and generate income.

This study identifies key factors that can potentially affect investors’ decision-making processes. First, clinical studies play a crucial role in drug development and contribute significantly to the overall cost. Challenges, such as limited clinical data from reference products and complexities associated with different types of IMDs, along with drug properties, such as pharmacodynamics, pharmacokinetics, and toxicity, can drive the investment required for this phase. Consequently, this cost burden can impact investors’ decisions and hinder the development of new drugs in the domestic pharmaceutical industry. When it is not feasible to bridge clinical study data from a previous product to support the development of a new drug, conducting a clinical study becomes imperative.(9) Offering incentives and support to conduct clinical studies, particularly for IMDs, can alleviate the financial burden on pharmaceutical companies. These incentives may include grants, tax benefits, or expedited regulatory processes. In such cases, an alternative strategy could involve considering clinical trials in countries with high overall attractiveness indices(11). In doing so, the aim is to expedite the drug’s path to the market, ultimately accelerating the return on investment and potentially gaining a competitive edge over other market players.

Second, the development of IMDs in domestic industry is still relatively new, resulting in longer development times and increased costs. One key factor contributing to this extended duration is the limited knowledge and expertise within the domestic industry and related organizations. Additionally, the conducting of clinical trials and challenges in the registration system can add to the overall time required for the development of an IMDs. To mitigate these challenges and shorten the duration of development, it is essential to focus on building capacity among all stakeholders involved in the process by establishing comprehensive capacity-building programs to enhance expertise within the domestic pharmaceutical industry and related organizations. These programs should focus on improving proficiency in the R&D processes, conducting efficient clinical trials, and navigating regulatory procedures. By implementing such initiatives, the duration of the IMDs development can be reduced, resulting in cost savings. Furthermore, it is crucial to recognize that the R&D process for IMDs should commence early during the originator-branded product lifecycle. Since IMDs development can be time-consuming, starting the R&D process early ensures sufficient time for thorough research, testing, and regulatory processes.

Third, owing to the high investment required to reach the capital break-even point and generate profits, entrepreneurs need to consider market feasibility and potential sales opportunities for IMDs. The presence of generic and new generic drugs as competitors in the market increases the complexity. To promote the adoption of IMDs, the government and associated organizations can play a role by proposing supportive policies as well as supporting export policies to expand market opportunities. Collaborative efforts involving the public and private sectors are crucial for sustainable development of the domestic pharmaceutical industry.

Fourth, the process of selecting suitable drugs for IMDs development involves careful consideration of various factors including financial aspects, patient needs, scientific and technological feasibility, and legal and registration feasibilities. It is crucial to acknowledge that not every drug can be formulated in every dosage form, which makes the drug selection process a significant factor that influences investment decisions. Several key elements are involved in drug selection. Market competitors and the expected payback period that the pharmaceutical industry can accept play crucial roles in determining the feasibility of drugs for IMD development. For instance, drugs such as antibiotics may require shorter payback periods owing to the potential emergence of drug resistance, whereas drugs used to treat chronic diseases or orphan drugs may require longer payback periods. In this context, selecting drugs with longer lifecycles and those that already possess sufficient existing data for bridging may be more feasible investment options. These drugs are more likely to have sustainable market demand and provide a higher chance of success in the development process.

By adopting and implementing these policy recommendations, the government and associated organizations can establish an enabling environment for the development of IMDs, foster innovation, reduce costs, and ensure long-term sustainability of the domestic pharmaceutical industry.

## Conclusions

This study provides crucial insights into the financial feasibility of developing sustained release dosage forms for IMDs in the Thai pharmaceutical industry. The proposed policy recommendations aim to create an enabling environment for IMDs development, ultimately fostering innovation and ensuring the sustainability of the domestic pharmaceutical industry.

## Data Availability

All data files are available from the figshare with this URL https://doi.org/10.6084/m9.figshare.25855492.v1

## Acknowledgements

The authors would like to express my sincere gratitude to the Thai Pharmaceutical Manufacturers Association (TPMA) for their excellent collaboration, which allowed us to conduct interviews and gather valuable data for the assessment. Our deepest appreciation goes to all the participants who generously gave their time and contributed to this study. Their active participation and valuable insights greatly enriched our research.

